# Light exposure during sleep is associated with irregular sleep timing: the Multi-Ethnic Study of Atherosclerosis (MESA)

**DOI:** 10.1101/2023.10.11.23296889

**Authors:** Danielle A Wallace, Xinye Qiu, Joel Schwartz, Tianyi Huang, Frank A.J.L. Scheer, Susan Redline, Tamar Sofer

## Abstract

**Objective:** Exposure to light at night (LAN) may influence sleep timing and regularity. Here, we test whether greater light exposure during sleep (LEDS) associates with greater irregularity in sleep onset timing in a large cohort of older adults.

**Methods:** Light exposure and activity patterns, measured via wrist-worn actigraphy (ActiWatch Spectrum), were analyzed in 1,933 participants with 6+ valid days of data in the Multi-Ethnic Study of Atherosclerosis (MESA) Exam 5 Sleep Study. Summary measures of LEDS averaged across nights were evaluated in linear and logistic regression analyses to test the association with standard deviation (SD) in sleep onset timing (continuous variable) and irregular sleep onset timing (SD≥1.36 hours, binary). Night-to-night associations between LEDS and absolute differences in nightly sleep onset timing were also evaluated with distributed lag non-linear models and mixed models.

**Results:** In between-individual linear and logistic models adjusted for demographic, health, and seasonal factors, every 5-lux unit increase in LEDS was associated with an increase of 7.8 minutes in sleep onset SD (β=0.13 hours, 95%CI:0.09-0.17) and 40% greater odds (OR=1.40, 95%CI:1.24-1.60) of irregular sleep onset. In within-individual night-to-night mixed model analyses, every 5-lux unit increase in LEDS the night prior (lag0) was associated with a 2.2-minute greater deviation of sleep onset the next night (β=0.036 hours, p<0.05). Conversely, every 1-hour increase in sleep deviation (lag0) was associated with a 0.35-lux increase in future LEDS (β=0.347 lux, p<0.05).

**Conclusion:** LEDS was associated with greater irregularity in sleep onset in between-individual analyses and subsequent deviation in sleep timing in within-individual analyses, supporting a role for LEDS in exacerbating irregular sleep onset timing. Greater deviation in sleep onset was also associated with greater future LEDS, suggesting a bidirectional relationship. Maintaining a dark sleeping environment and preventing LEDS may promote sleep regularity and following a regular sleep schedule may limit LEDS.

## 1. Introduction

Sleep timing is regulated by two primary biological processes: 1) the homeostatic sleep drive and 2) a circadian rhythm in sleep propensity^1^. For the circadian regulation of sleep timing, light is the most important zeitgeber, or “time giver”^2^, and in this way, light can contribute to stability or irregularity in sleep-wake timing. Controlled laboratory studies have shown that light exposure shifts circadian rhythms in an intensity-, wavelength-, light history-, and duration-dependent manner to advance or delay the phase of circadian rhythms, dependent on the timing of exposure^2,3^. These studies have shown that the human circadian system is most sensitive to the phase-shifting effects of light during the “biological” night^3,4^, the time just preceding, during, and following the habitual sleep episode. Light exposure in the earlier part of the biological night delays (moves later), whereas light exposure in the latter half of the night advances (moves earlier), the circadian pacemaker^3^, a cluster of neurons in the suprachiasmatic nucleus of the hypothalamus that autonomously keeps time and generates timing signals for other body regions^5^. Individuals may differ in sensitivity to the phase-shifting effects of light at night (LAN)^6^. Whether light advances or delays sleep timing, and by how much, depends on the endogenous circadian phase during light exposure^2^. In addition to the phase-shifting effects of light, light also has acute negative effects on sleep propensity, in part through suppression of melatonin^7^.

Our modern light environment is drastically different from the one in which humans evolved, where sunlight provided light during the day and (much dimmer) moonlight or fires were the only nighttime light sources. These natural LAN exposures had a limited impact on the circadian system in comparison with modern electric lighting^8^. In comparison, people now have access to inexpensive, abundant, and bright light at any time of day or night with modern electrical lighting^9,10^. Because the phase-shifting effects of light on the human circadian timing system are strongest around the biological night, exposure to LAN in the home environment may influence the circadian system. Therefore, in addition to the acute effects that LAN can have on sleep, LAN may also influence sleep timing. However, epidemiological studies of sleep in naturalistic settings may not have measures of circadian phase that could be used to model the phase-shifting effects of light. In such cases, measuring light exposure during sleep (LEDS) could provide an alternative way to capture the effects of light exposure during a biologically sensitive window.

A particularly important dimension of sleep timing is its nightly regularity or variability. Irregular sleep timing, defined as variability in night-to-night sleep timing, may represent a type of circadian misalignment common in the general population^11^. Greater irregularity in sleep timing has been linked to mechanisms for chronic disease^12^, such as increased inflammatory biomarkers^13^, and metabolic syndrome^14^. Experimental sleep restriction studies with and without circadian disruption further support that circadian misalignment itself has adverse cardiometabolic consequences above and beyond any effects of sleep restriction^15–18^. In particular, timing and irregularity of sleep onset time have been associated with increased cardiovascular disease risk^19,20^ and abnormal metabolic markers^14^.

LAN and LEDS are potentially modifiable lifestyle risk factors. Because reverse causation may exist for testing the effects of naturalistic pre-sleep light exposure on the sleep episode (e.g., people with irregular sleep schedules may have later chronotypes and be more active at night), the evaluation of LEDS rather than pre-sleep light may limit this bias. If LEDS is greater in people with irregular sleep timing, reducing LEDS may present an opportunity to establish more regular sleep patterns and improve downstream health outcomes. Because the interplay of environmental light exposure and sleep timing regularity in the general population has not been well characterized^21^, here, we tested the hypothesis that greater LEDS is associated with less regularity in sleep onset timing in a large, multiethnic cohort studied in their usual home environments.

## 2. Methods

### 2.1 Study population

This analysis utilized data from the Multi-Ethnic Study of Atherosclerosis (MESA), designed to study subclinical and clinical cardiovascular disease in a multiracial, multiethnic sample of adults 45-84 years of age at study initiation. The baseline Exam 1 study (2000-2002) enrolled 6,814 participants at six U.S. study sites (Los Angeles, CA; St. Paul, MN; Chicago, IL; Forsyth County, NC; Baltimore, MD; and Manhattan and Southern Bronx, NY)^22^. To be initially included in the cohort, participants had to be free of known cardiovascular disease or other health conditions that would prevent follow up examination. Subsequent examinations of the cohort were completed over approximately 20 years. Between 2010-2013, actigraphy data that included both sleep-wake and light measurements were collected as part of the MESA Exam 5 Sleep Ancillary study (n=2,261). Of the 2,159 participants with actigraphy and light records, we excluded participants with <6 valid nights. The analysis included demographic and health data as covariates from study questionnaires. The institutional review board at each study site approved the study, and all participants provided written informed consent.

### 2.2 Actigraphy data processing and derivation of light exposure during sleep (LEDS)

Objective sleep and white light data were collected in 30-second epochs with ActiWatch Spectrum actigraphy devices (Philips Respironics, Murrysville, PA; wavelength sensitivity 400 – 700nm) worn on the non-dominant wrist. Sleep actigraphy data were scored by trained personnel at Brigham and Women’s Hospital Sleep Reading Center^23^. Sleep onset for the main sleep episode and nap times were annotated based on hierarchical methods utilizing ActiWatch event marker, participant sleep journal, light levels, and/or activity count data, as previously described^24^. Measures of light exposure, sleep timing, behavioral activity, and sleep duration were derived for participants with at least 6 valid nights (main sleep episode) of data from the first 7 consecutive days of actigraphy data. Sleep-wake was determined using the Philips Actiware 5.59 algorithm with the intermediate (40 counts per epoch) setting for wake detection, with sleep onset defined as 5-minute period of immobility. Timeseries data for each participant were processed to define a person-specific “day” using the sleep onset timing for the first sleep episode, rather than midnight, as the day start time. Consequently, the first sleep episode began at the beginning of the person-specific day. All daily measures described below refer to the person-specific day. Using this definition of a day, average behavioral activity, average white light (lux) exposure during sleep (LEDS), and the sleep episode duration were calculated. If there were >10 cumulative minutes of missing white light or activity data during the main sleep episode, or if the main sleep episode lasted <2 hours or >14 hours, the night was considered invalid and that night’s LEDS and sleep duration were set to missing. White light lux values within the main sleep episode (from sleep onset to sleep offset) were averaged for a mean nightly LEDS value. Additionally, participants with an average LEDS value (across all nights) higher than the 99^th^ percentile were excluded.

### 2.3 Derivation of sleep regularity outcome variables

The outcome for the analyses of sleep regularity was SD of sleep onset timing across nights. To calculate sleep onset timing SD, the circular standard deviation (SD)^25^ of the timestamps across each night’s sleep onset was calculated. Sleep onset, rather than other markers of sleep timing such as sleep midpoint or sleep offset, was chosen as the outcome because of its relevance as a marker of the circadian gating of the wake-maintenance zone and propensity for sleep^26^ and ease of interpretability. Additionally, the temporality of measures ensures that sleep onset occurs prior to the LEDS measure for that sleep episode, while still allowing LEDS to influence sleep onset timing. The outcomes for the night-to-night distributed lag nonlinear model (DLNM) and mixed model analyses were the absolute circular difference or deviation in sleep onset timing from night to night.

### 2.4 Covariates

Covariates were chosen after review of the literature. Race and ethnicity are social constructs; the self-reported MESA-defined groups of race/ethnicity (see categories below) were included as a covariate to attempt to account for the influence of racism and structural bias on sleep. The term “gender” is used here to reflect the wording used in MESA questionnaires, after which participants were asked to indicate “Male” or “Female”, but we acknowledge that this variable may reflect sex and/or not appropriately capture gender identity. Other covariates included: age (years) during the Sleep Ancillary Exam, self-reported gender from Exam 1 (male/female), self-reported race and ethnicity from Exam 1 using MESA-defined groups (Black, Chinese, Hispanic/Latino, or White), Exam 5 employment status (“no” (ref) or “yes”), partner status (married or living with a partner (ref) vs other), and federal poverty level status, a binary variable derived from self-reported income and household size to reflect 2010-2012 federal poverty level status (above or below federal poverty level). Clinical site location and season were also included; season derived from the month of the first day of actigraphy measurement, with cut points based the solstice and equinox dates: November-January, February-April, May-July, August-October. Health-behavior related covariates included: smoking status at Exam 5 (current=”yes”), chronotype (summary score, modeled as continuous), waist-to-hip ratio (WHR^27^, continuous), and average behavioral activity (continuous). Self-reported chronotype was measured using the modified Horne-Ostberg Morningness-Eveningness Questionnaire score^28^ (higher scores indicate greater morningness) during the ancillary sleep study during Exam 5. Waist and hip circumference were measured during Exam 5. Daily average behavioral activity counts (across both wake and sleep episodes) were calculated from accelerometer activity counts from wrist actigraphs and averaged across person-specific days as an objective measure of behavioral activity (continuous); if >8 cumulative hours of actigraphy data were missing across the day, that day’s average activity was set to missing. Sleep duration was calculated as the nightly duration (hours) of the main sleep episode interval and sleep fragmentation index was calculated as the proportion of immobile to mobile epochs^24^. For the sensitivity analysis excluding shift workers, work schedules were ascertained during Exam 5 with the question “Which of the following best describes your usual work schedule?” (options: Day/Afternoon/Night/Split/Irregular/Rotating/Don’t work) and participants who responded with a shift other than “Day” or “Don’t work” were considered shift workers. For the sensitivity analysis excluding participants with insomnia, people who responded “yes” during Exam 5 to the question “Have you ever been told by a doctor that you have any of the following?” with “Insomnia?” listed (options: No/Yes) and/or people with a Women’s Health Initiative Insomnia Rating (WHIIR) score of 9 or higher^29^ were excluded.

### 2.5 Statistical analyses

In the primary between-individual linear and logistic analyses, sleep onset SD was analyzed as a continuous and dichotomous outcome with LEDS (averaged across nights) as the exposure of interest while adjusting for covariates in Models 1-5. Model 1 adjusted for age, gender, and race and ethnicity. Model 2 additionally adjusted for employment status, partner status, and federal poverty level status. Model 3 additionally adjusted for smoking, chronotype, WHR, and average behavioral activity. Model 4 additionally adjusted for the site and season; results from Model 4 are presented as the main results in the text. Lastly, average sleep duration and sleep fragmentation were identified as possible confounders (which are also possible colliders) and modelled as continuous covariates in Model 5. Sleep duration and sleep fragmentation index were modelled as averaged values in the logistic and lag analyses and as nightly measures in the mixed model analyses.

LEDS was modeled both as a continuous and categorical variable (tertiles), with the lower limit of the third tertile aligning closely with the 1 lux benchmark for sleep environment recommendations^30^. Sleep onset SD had a right-skewed distribution and was also modeled as a continuous variable after Box-Cox transformation using the “MASS” package^31^ (**Supplemental Material**). Sleep onset SD was also modeled as a dichotomized outcome in logistic regression models, with “regular” sleep timing defined as individuals in the bottom 75% sleep onset SD values and “irregular” sleep timing as individuals in the top 25% sleep onset SD values (cutoff = 1.36 hours SD).

Because the effect of LEDS on sleep timing may have delayed or non-linear effects, secondary analyses modeled the night-to-night lagged associations of LEDS (e.g., LEDS night 5=lag0, LEDS night 4=lag-1, LEDS night 3=lag-2, LEDS night 2=lag-3, LEDS night 1=lag-4) on absolute deviation in sleep onset timing from night 5 to night 6 (circular difference) to identify the optimal window for the exposure-outcome relationship. Lags are denoted with a “-“ to represent going back in time in relation to the modelled outcome. DLNM models were constructed using the “dlnm” package^32^. Deviation in sleep onset timing was also modeled as the predictor and LEDS (night 6) as the outcome. Associations were estimated with generalized additive models with penalized splines^33^. The exposure-response function was modeled as a natural spline and the lag-response function as a penalized spline with internal knots selected based on smallest AIC (details in **Supplemental Material**).

Using the lag period(s) identified with DLNM modeling, night-to-night within-individual associations between LEDS and irregularity in sleep onset timing were further modeled using mixed linear regression modeling. The absolute deviation in sleep timing from night 1 to night 2, night 2 to night 3, night 3 to night 4, night 4 to night 5, and night 5 to night 6 were modeled as the repeated-measure dependent variable, while LEDS during night 1, night 2, night 3, night 4, and night 5 were modeled as the repeated-measure exposure. In Model 5, nightly sleep duration and sleep fragmentation index (nightly measures from nights 1-5 instead of averaged values) were included as fixed effects. To explore bidirectionality, deviation in sleep onset (nights 1-2, 2-3, 3-4, 4-5, 5-6) was also modeled as the exposure and LEDS (nights 2, 3, 4, 5, 6) modeled as the dependent variable. The covariance structure was specified as first-order autoregressive structure with heterogenous variances (correlation=corAR1(form=∼1|ID)) and ID modeled as a random effect to allow for variability between participants using the “nlme” package^34^ in R. A sensitivity analysis excluding shift workers and a sensitivity analysis excluding people with insomnia were also conducted for the logistic and mixed model regressions. All statistical analyses were performed in R version 4.1.1. and results considered statistically significant if p-value <0.05.

## 3. Results

### 3.1 Demographic characteristics

After excluding 206 participants with <6 valid nights of data, 1,933 MESA participants were included in the analysis (**Supplemental Table 1**). A representation of the study design is provided in **Figure 1**. On average, participants were 69 years of age, 55% female, and the majority were retired or unemployed. The average sleep onset timing among the sample was approximately 11:35PM (median=10:19PM, sd=1.37 hours). The range in LEDS at the 1^st^, 2^nd^, and 3^rd^ tertiles were 0-0.33, 0.33-0.95, and 0.95-39.4 lux, and the range in sleep onset SD at the 1^st^, 2^nd^, 3^rd^ and 4^th^ quartiles were 0.06-0.56 hours, 0.56-0.90 hours, 0.90-1.35 hours, and 1.35-6.9 hours, respectively. Approximately 43% of the sample had a sleep onset variability (SD) of 1 hour or more and 31.5% had an average LEDS value of 1 lux or brighter (**Supplemental Table 2**). Overall, participants in the highest tertile of LEDS exposure (≥0.95 lux) were older, had a lower MEQ score, and differed by season and site (**Table 1**). Greater LEDS exposure was also more common among Black participants and less common among Hispanic/Latino participants (**Table 1**). Visual inspection of averaged light levels stratified by sleep regularity group suggested greater LEDS among irregular sleepers (≥1.36 hours SD; top quartile) compared to regular sleepers (**Figure 2**).

**Figure 1.**
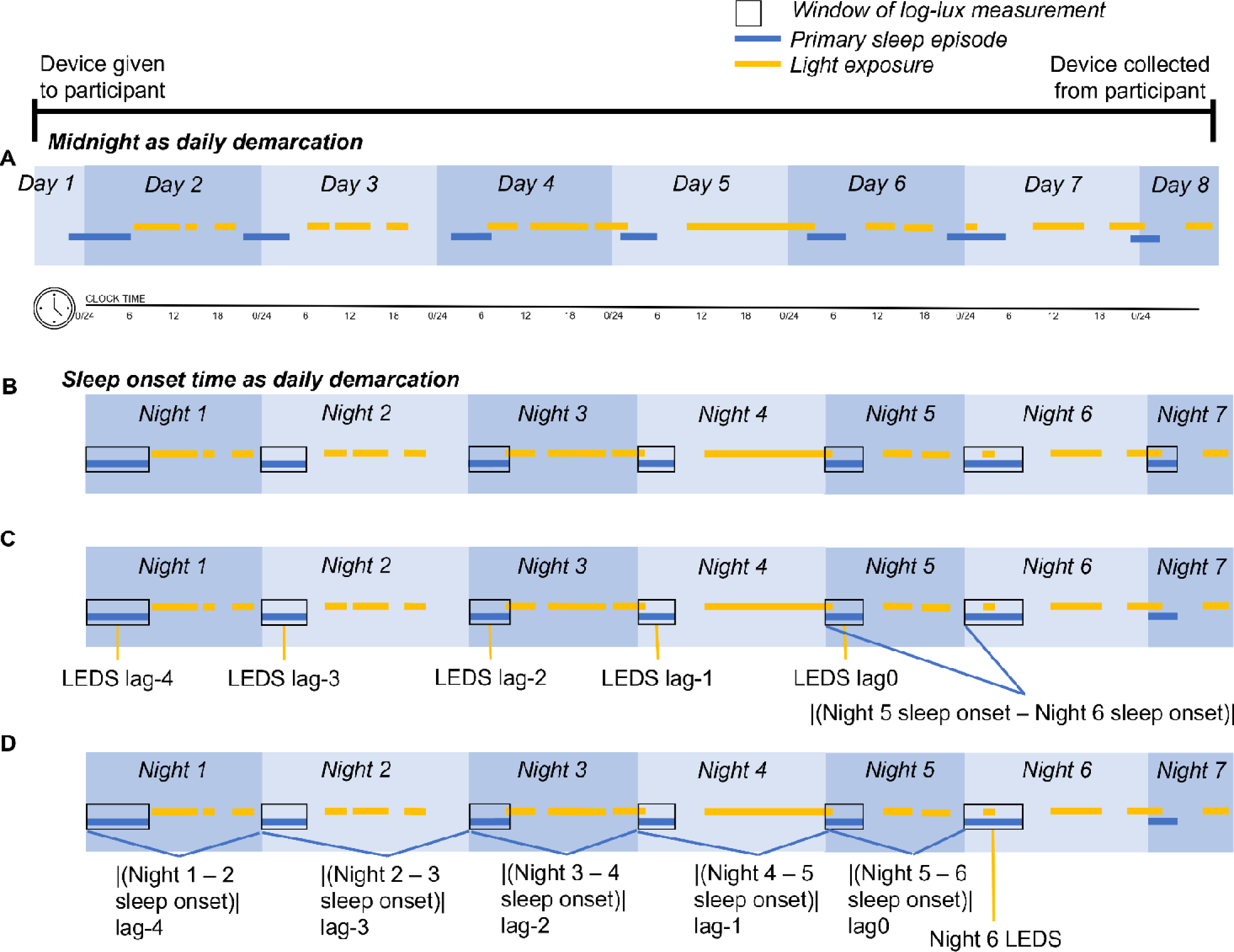
Depiction of example data timeline and study design. Horizontal yellow bars indicate light exposure and horizontal blue bars indicate the sleep episode; black boxes indicate the window of the sleep episode during which nightly average light exposure during sleep (LEDS) was calculated. Rather than using (**A**) midnight to demarcate daily measures, this analysis used (**B**) sleep onset timing to demarcate daily measures. (**B**) Logistic regression analyses analyzed data averaged across days, whereas (**C**) the night-to-night analysis distributed lag non-linear model (DLNM) analyzed LEDS on the 5^th^ night (lag0), the 4^th^ night (lag-1), the 3^rd^ night (lag-2), the 2^nd^ night (lag3-) and the 1^st^ night (lag-4) as the exposure and the absolute difference in sleep onset timing between the 5^th^ and 6^th^ night as the outcome. Likewise, (**D**) another DLNM analysis modelled the absolute difference in sleep onset timing between the 5^th^ and 6^th^ nights (lag0), the 4^th^ and 5^th^ nights (lag-1), the 3^rd^ and 4^th^ nights (lag-2), and 2^nd^ and 3^rd^ nights (lag-3), and the 1^st^ and 2^nd^ nights (lag-4) as the exposure and LEDS on the 6^th^ night as the outcome. Night to night measures were further analyzed in mixed models using the lag identified in the DLNM.

**Figure 2.**
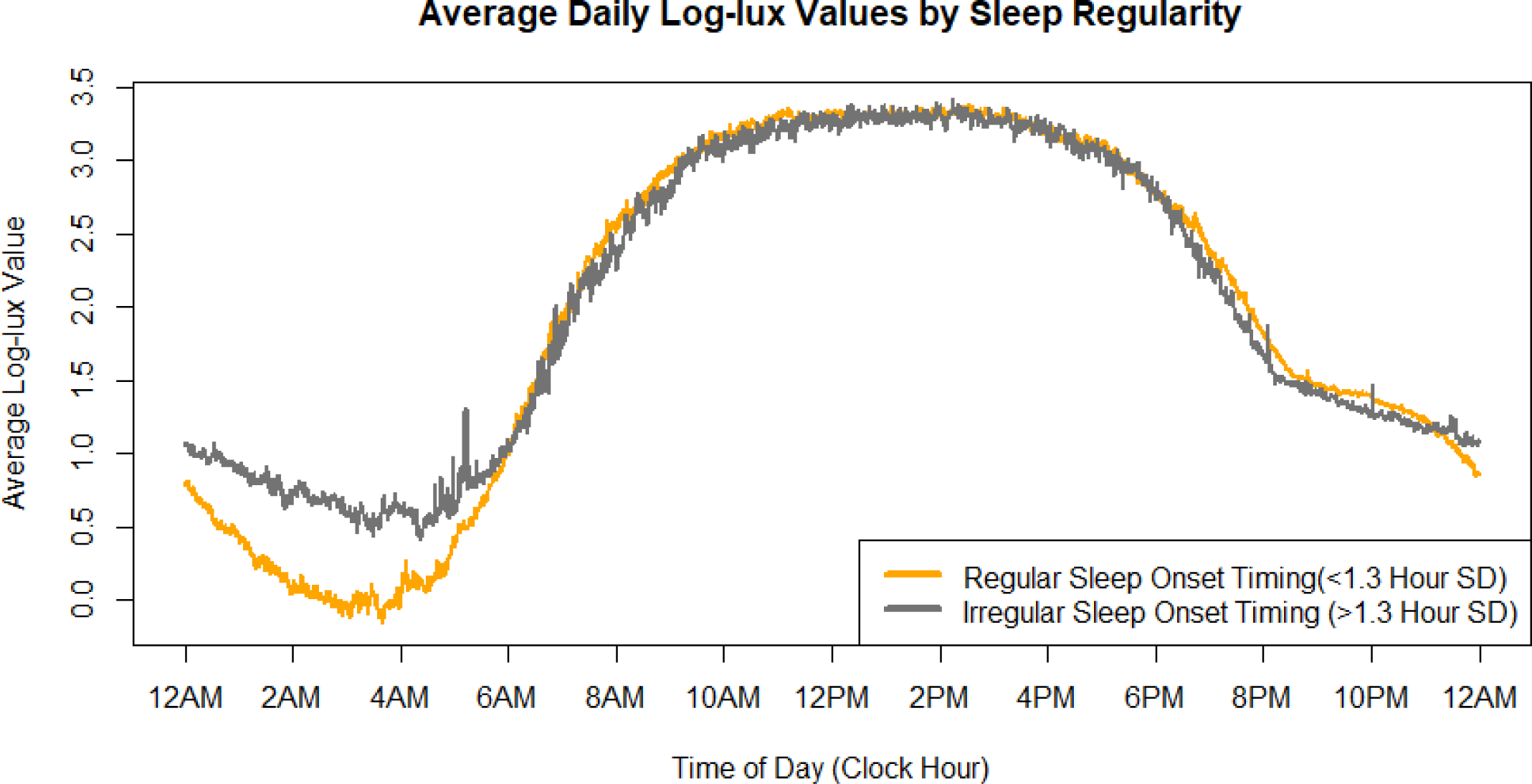
Average daily log10-lux illuminance values in MESA by time of day, stratified by the regular sleep onset timing group (n=1,459 participants) and the irregular sleep onset timing group (n=474 participants).

**Table 1.** Descriptive statistics of the study sample, stratified by LEDS tertiles.

|  |  |  |  |
| --- | --- | --- | --- |
| **Variables** | **LEDS Tertiles** |  |  |
|  | 0-0.33 lux<br>(n=645) | 0.33-0.95 lux<br>(n=644) | ≥0.95 lux<br>(n=644) |
| Age, years (mean (SD)) | 68.77 (9.09) | 68.95 (8.81) | 70.76 (9.39) |
| WHR (mean (SD)) | 0.93 (0.08) | 0.94 (0.08) | 0.94 (0.09) |
| Gender (N (% Male)) | 282 (43.7) | 299 (46.4) | 297 (46.1) |
| Race/ethnicity (N (%)) |  |  |  |
| *Chinese* | 68 (10.5) | 72 (11.2) | 79 (12.3) |
| *Hispanic/Latino* | 182 (28.2) | 160 (24.8) | 118 (18.3) |
| *Black* | 131 (20.3) | 178 (27.6) | 211 (32.8) |
| *White* | 264 (40.9) | 234 (36.3) | 236 (36.6) |
| Currently employed (N (% Yes)) | 154 (24.0) | 153 (23.9) | 155 (24.2) |
| Income below federal poverty (N (% Yes)) | 45 (7.2) | 46 (7.4) | 41 (6.6) |
| Partner status, married or currently living with partner (N (% Yes)) | 389 (61.3) | 402 (63.3) | 368 (58.0) |
| Current smoking (N (% Yes)) | 43 (6.7) | 46 (7.2) | 36 (5.6) |
| Chronotype MEQ score (mean (SD)) | 17.60 (3.48) | 17.58 (3.49) | 16.61 (3.77) |
| Season of measurement |  |  |  |
| *August-October* | 139 (21.6) | 168 (26.1) | 198 (30.7) |
| *November-January* | 180 (27.9) | 137 (21.3) | 116 (18.0) |
| *February-April* | 200 (31.0) | 169 (26.2) | 117 (18.2) |
| *May-July* | 126 (19.5) | 170 (26.4) | 213 (33.1) |
| Clinical Site (N (%)) |  |  |  |
| *Wake Forest University* | 108 (16.7) | 105 (16.3) | 95 (14.8) |
| *Columbia University* | 126 (19.5) | 92 (14.3) | 125 (19.4) |
| *Johns Hopkins University* | 72 (11.2) | 105 (16.3) | 106 (16.5) |
| *University of Minnesota* | 160 (24.8) | 125 (19.4) | 79 (12.3) |
| *Northwestern University* | 90 (14.0) | 113 (17.5) | 133 (20.7) |
| *University of California, Los Angeles* | 89 (13.8) | 104 (16.1) | 106 (16.5) |
| Shiftwork (N (% Yes)) | 75 (11.6) | 77 (12.0) | 83 (12.9) |
| Insomnia (N (% Yes)) | 236 (36.6) | 237 (36.8) | 244 (37.9) |
| Average LEDS, mean (SD) | 0.14 (0.10) | 0.58 (0.17) | 4.65 (6.23) |

### 3.2 LEDS averaged across days is associated with irregular sleep onset timing

In the primary linear and logistic regression between-individual analyses, greater average LEDS was associated with greater irregularity in sleep timing (**Tables 2-3**). When sleep timing variability was modeled as a continuous outcome, every 1-lux and 5-lux unit increase was associated with a 1.8-minute (β=0.03 hours, 95% CI: 0.02, 0.03) and 7.8-minute (β=0.13 hours, 95% CI: 0.09, 0.17) increase in sleep onset SD, respectively (**Table 2**, Model 4 results). When analyzed as tertiles, the highest LEDS tertile (≥0.95 lux) was associated with a 13.2-minute increase in sleep onset SD (β=0.22 hours, 95% CI: 0.14, 0.30). Results from linear models with Box-Cox transformed sleep onset SD were similar (**Supplemental Figure 1, Supplemental Table 3**). Likewise, when sleep timing variability was dichotomized and analyzed in logistic regression, every 1-lux and 5-lux unit increase was associated with approximately 7% (OR=1.07, 95% CI: 1.04, 1.10) and 40% (OR=1.40, 95% CI: 1.24, 1.60) higher odds of irregular (≥1.36 hours SD) sleep timing, respectively (**Table 3**, Model 4 results). This association decreased to approximately 6% and 36% higher odds of irregular sleep timing when average sleep duration and the sleep fragmentation index were included. When analyzing LEDS as tertiles, those in the highest exposure group had 72% (OR=1.72, 95% CI: 1.31, 2.28) higher odds of irregular sleep timing compared to the lowest tertile (**Table 3**).

**Table 2.** Linear regression results for light exposure during sleep (LEDS, continuous or tertiles) with sleep onset timing variability (SD, hours) as the outcome.

|  | Crude<br>[95% CI] | Model 1<br>[95% CI] | Model 2<br>[95% CI] | Model 3<br>[95% CI] | **Model 4**<br>**[95% CI]** | Model 5<br>[95% CI] |
| --- | --- | --- | --- | --- | --- | --- |
| *LEDS as continuous\*:* |  |  |  |  |  |  |
| *LEDS (per 1-unit lux)* | **0.03**<br>(0.02-0.04) | **0.03**<br>(0.02-0.04) | **0.03**<br>(0.02-0.03) | **0.03**<br>(0.02-0.03) | **0.03**<br>(0.02-0.03) | **0.02**<br>(0.02-0.03) |
| *LEDS as tertiles\*:* |  |  |  |  |  |  |
| *LEDS T1 (0-0.33 lux)* | (ref) | (ref) | (ref) | (ref) | (ref) | (ref) |
| *LEDS T2 (0.33-0.95 lux)* | 0.01<br>(-0.07-0.08) | -0.01<br>(-0.09-0.06) | -0.02<br>(-0.09-0.06) | -0.01<br>(-0.08-0.07) | -0.01<br>(-0.08-0.07) | 0.02 (-0.06-0.09) |
| *LEDS T3 (0.95-39.4 lux)* | **0.25**<br>(0.17-0.32) | **0.23**<br>(0.16-0.30) | **0.23**<br>(0.16-0.31) | **0.22**<br>(0.15-0.30) | **0.22**<br>(0.14-0.30) | **0.20**<br>(0.13-0.27) |

\*Sleep onset SD (hours) as outcome
Crude and adjusted linear regression model estimates with 95% confidence intervals are presented, with $p < 0.05$ in bold. Model 1 adjusted for age, gender, and race/ethnicity; Model 2 adjusted for all the covariates included in Model 1 in addition to poverty, employment status, and partner status; Model 3 adjusted for all the covariates included in Model 2 in addition to smoking, chronotype, waist to hip ratio, and average behavioral activity; Model 4 adjusted for all the covariates included in Model 3 in addition to season and Exam 5 site; Model 5 adjusted for all the covariates in Model 4 in addition to average sleep episode duration, and average sleep fragmentation index.

**Table 3.** Logistic regression results for light exposure during sleep (LEDS, continuous or tertiles) with irregular sleep onset timing (approximately ≥ 1.36 hours SD).

|  | Crude<br>[95% CI] | Model 1<br>[95% CI] | Model 2<br>[95% CI] | Model 3<br>[95% CI] | **Model 4**<br>**[95% CI]** | Model 5<br>[95% CI] |
| --- | --- | --- | --- | --- | --- | --- |
| *LEDS as continuous\*:* |  |  |  |  |  |  |
| *LEDS (per 1-unit lux)* | **1.08**<br>(1.05-1.10) | **1.08**<br>(1.05-1.10) | **1.08**<br>(1.05-1.10) | **1.07**<br>(1.05-1.10) | **1.07**<br>(1.04-1.10) | **1.06**<br>(1.04-1.09) |
| *LEDS as tertiles\*:* |  |  |  |  |  |  |
| *LEDS T1 (0-0.33 lux)* | (ref) | (ref) | (ref) | (ref) | (ref) | (ref) |
| *LEDS T2 (0.33-0.95 lux)* | 1.04<br>(0.80-1.36) | 0.97<br>(0.74-1.28) | 0.95<br>(0.72-1.26) | 0.98<br>(0.74-1.30) | 0.98<br>(0.73-1.30) | 1.04<br>(0.77-1.40) |
| *LEDS T3 (0.95-39.4 lux)* | **1.86**<br>(1.45-2.39) | **1.74**<br>(1.35-2.26) | **1.75**<br>(1.34-2.28) | **1.76**<br>(1.34-2.31) | **1.72**<br>(1.31-2.28) | **1.66**<br>(1.24-2.21) |

Crude and adjusted odds ratios (ORs) with 95% confidence intervals are presented. ORs with $p < 0.05$ are in bold. Model 1 adjusted for age, gender, and race/ethnicity; Model 2 adjusted for all the covariates included in Model 1 in addition to poverty, employment status, and partner status; Model 3 adjusted for all the covariates included in Model 2 in addition to smoking, chronotype, waist to hip ratio, and average behavioral activity; Model 4 adjusted for all the covariates included in Model 3 in addition to season and Exam 5 site; Model 5 adjusted for all the covariates in Model 4 in addition to average sleep episode duration, and average sleep fragmentation index.

### 3.3 LEDS up to two nights prior is associated with future sleep onset timing deviation in lagged analyses

In the secondary analyses, to identify the appropriate lag for the exposure-response relationship, we investigated night-to-night lagged associations between LEDS (predictor) and deviation in sleep onset timing (night 5 to night 6 as the outcome, **Figure 1C**). The single-night lag-response curves for predicted LEDS on sleep timing are shown in **Supplemental Figure 2**, with a positive association between greater lux and greater deviation in sleep timing. Increasing LEDS from 0 to 0.5 lux was predicted to increase sleep deviation by 0.6 minutes at lag0 (β=0.01 hours; 95% CI:0.002,0.018; **Supplemental Figure 2A**) and 0.3 minutes at lag-1 (β=0.005 hours; 95% CI:0.0004, 0.01), but not at other lags. Likewise, increasing LEDS from 0 to 1 lux was predicted to increase deviation in sleep timing by 1.1 minutes at lag0 (β=0.019 hours; 95% CI:0.003,0.036; **Supplemental Figure 2B**) and 0.6 minutes at lag-1 (β=0.01 hours; 95% CI:0.0007, 0.02). Greater increases from 0 to 3 lux or 0 to 10 lux were associated with deviations of 3.4 and 10.7 minutes at lag0, respectively (β=0.057 hours; 95% CI:0.009, 0.105; β=0.178 hours; 95% CI:0.031, 0.325; **Supplemental Figure 2C-D**), and 1.8 and 5.5 minutes at lag-1 (β=0.03 hours; 95% CI:0.002, 0.06; β=0.09 hours; 95% CI:0.007, 0.177). However, other preceding timepoints (lag-2 to lag-4) were not associated. The exposure-outcome response surface had a saddle shape, with the greatest slope at lag0 (**Supplemental Figure 2E**). **Supplemental Figure 2F** shows the cumulative influence of LEDS on sleep deviation (across lags), with confidence bands becoming progressively wider as lux increases.

### 3.4 Night-to-night within-individual analysis suggests LEDS affects future sleep onset timing deviation

Repeated nightly measures of LEDS and absolute sleep onset deviation were next analyzed within individuals with mixed linear models using the lag window identified by the DLNM analysis (lag0). LEDS was significantly associated with deviation in sleep onset the following night, with or without adjusting for that night’s sleep fragmentation and sleep duration. Every 1-lux and 5-lux increase in nightly LEDS was associated with approximately 0.42 minutes (β=0.007 hours, p<0.05; Model 4) and 2.2 minutes, respectively, increase in sleep deviation the following night (**Table 4**).

**Table 4.**
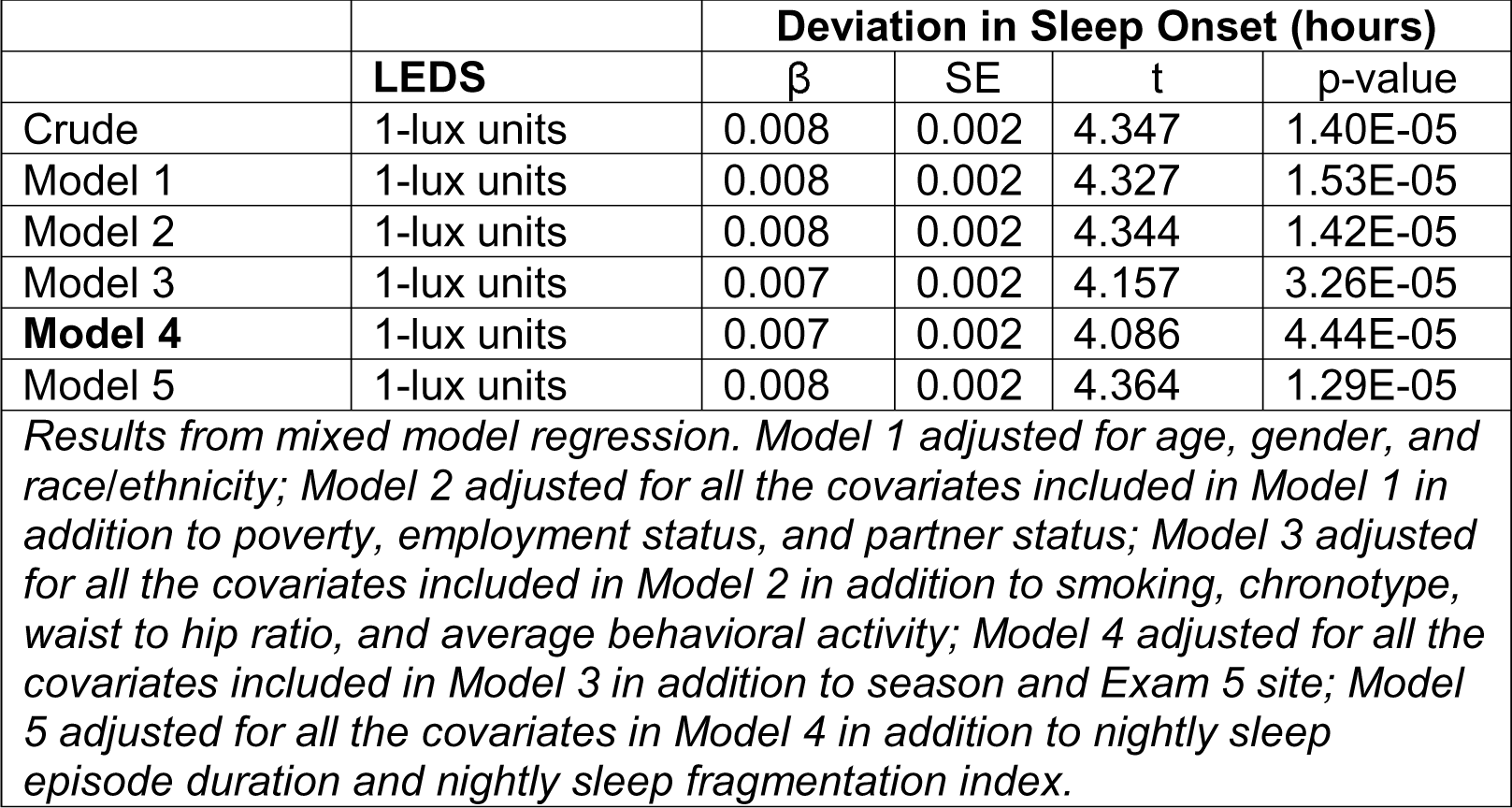
Results of mixed model regression for night-to-night associations between LEDS (exposure) and absolute deviation in sleep onset (outcome).

### 3.5 Large deviations in sleep timing the night before is associated with LEDS the following night in lagged analyses

A secondary lagged analysis modelled the lagged effects of sleep onset timing deviation (predictor, night 5 to 6=lag0, night 4 to 5=lag-1, etc.) on LEDS (outcome, night 6; **Figure 1D**). The single-night lag-response curves did not show associations between sleep deviation and LEDS at lag0 until sleep timing deviation became 4 hours or greater (**Supplemental Figures 3A-C**, **Supplemental Table 2**). A large deviation of 4 or 5 hours was predicted to increase LEDS (β=1.059 lux; 95% CI:0.248, 1.870; β=1.598 lux; 95% CI:0.704, 2.492; **Supplemental Figure 3D**). However, deviations of 1-3 hours showed some associations at longer lags (lag-2 to lag-4); for example, a 2 hour deviation a few nights prior (lag-2 to lag-4), but not the night or two before (lag0 to lag-1), was predicted to increase night 6 LEDS by 0.46 lux (lag-2), 0.55 lux (lag-3), and 0.64 lux (lag-4), respectively (β=0.464 lux; 95% CI:0.186, 0.743; β=0.552 lux; 95% CI:0.163, 0.941; β=0.640 lux; 95% CI:0.032, 1.247). These inconsistencies may be due to underlying variation in the data, as shown in the curve of the exposure-outcome response surface (**Supplemental Figure 3E**). **Supplemental Figure 3F** shows the cumulative influence of sleep deviation on LEDS, with increasing effects and confidence bands widening after approximately 6 hours deviation.

### 3.6 Night-to-night within-individual analysis also suggests sleep onset timing deviation affects future LEDS

To evaluate whether there may be bidirectional effects whereby deviation in sleep onset timing may also affect LEDS, repeated nightly measures of absolute sleep onset deviation were next analyzed within individuals with mixed linear models using the same lag window (lag0) as the prior mixed model analysis. Deviation in sleep onset was significantly associated with LEDS the following night, with or without adjusting for sleep fragmentation and sleep duration. Every 1-hour increase in sleep timing deviation was associated with approximately 0.35 greater LEDS (p<0.05; Model 4) the following night (**Table 5**).

**Table 5.** Results of mixed model regression for night-to-night associations between absolute deviation in sleep onset (exposure) and LEDS (outcome).

|  | Deviation | LEDS (lux) |  |  |  |
| --- | --- | --- | --- | --- | --- |
| | | $\beta$ | SE | t | p-value |
| Crude | 1-hour units | 0.369 | 0.063 | 5.888 | 4.09E-09 |
| Model 1 | 1-hour units | 0.364 | 0.063 | 5.788 | 7.44E-09 |
| Model 2 | 1-hour units | 0.364 | 0.063 | 5.776 | 7.97E-09 |
| Model 3 | 1-hour units | 0.351 | 0.063 | 5.560 | 2.79E-08 |
| **Model 4** | 1-hour units | 0.346 | 0.063 | 5.484 | 4.30E-08 |
| Model 5 | 1-hour units | 0.347 | 0.063 | 5.498 | 3.98E-08 |

Results from mixed model regression. Model 1 adjusted for age, gender, and race/ethnicity; Model 2 adjusted for all the covariates included in Model 1 in addition to poverty, employment status, and partner status; Model 3 adjusted for all the covariates included in Model 2 in addition to smoking, chronotype, waist to hip ratio, and average behavioral activity; Model 4 adjusted for all the covariates included in Model 3 in addition to season and Exam 5 site; Model 5 adjusted for all the covariates in Model 4 in addition to nightly sleep episode duration and nightly sleep fragmentation index.

### 3.7 Excluding shift workers or participants with insomnia does not abrogate the association of LEDS or sleep timing deviation

To evaluate the influence of shift work on the measured outcomes, we conducted sensitivity analyses excluding the 235 participants who reported working an afternoon, night, split, irregular/on-call, or rotating shift. The timeseries plot was similar, although the gap between regular and irregular sleepers became smaller (**Supplemental Figure 4**). The logistic regression results remained largely the same, if slightly attenuated in a few of the models (**Supplemental Table 4**). Regression estimates were also largely unchanged in the night-to-night mixed model regression results (**Supplemental Table 5**).

Because sleep onset timing may be less accurately determined and the frequency of wake episodes during sleep may be greater in participants with insomnia^35^, we also conducted a sensitivity analysis excluding the 717 participants with a history of insomnia and/or with a WHIIR score ≥ 9. Timeseries plots of the log10-transformed averaged lux values did not substantially change (**Supplemental Figure 5**). Overall, logistic regression results had slightly greater estimates for the effects of LEDS on sleep irregularity among the 1,116 participants without insomnia (**Supplemental Table 6**). However, regression estimates were somewhat attenuated in the night-to-night mixed model regression results, decreasing from an estimate of 0.007 hours to 0.006 hours (β=0.006 hours, p<0.05; Model 4; **Supplemental Table 7**).

In the night-to-night mixed model regression analyses modeling the effect of sleep onset deviation on LEDS the following night, excluding shift workers led to attenuated effect estimates, decreasing from 0.35 to 0.20 (p<0.05; Model 4; **Supplemental Table 8**). However, excluding participants with insomnia led to greater effect estimates, increasing from 0.35 to 0.54 (p<0.05; Model 4; **Supplemental Table 9**).

## 4. Discussion

In this large sample of adults from 6 geographically diverse communities across the U.S., studied with multi-day assessments of sleep-wake patterns and ambient light exposure, we found a positive association between greater light exposure during the main sleep episode (LEDS) and greater irregularity in sleep onset timing. While recent lighting guidelines for chronobiological health recommend a sleeping environment that is darker than 1 lux illuminance^30^, 31.5% of our sample of older adults had an average illuminance during sleep above this threshold, suggesting the pervasiveness of suboptimal light exposure during sleep. Averaged across nights, greater LEDS was associated with greater sleep onset irregularity. In repeated measures night-to-night analyses, greater LEDS was associated with greater deviation in sleep onset timing the next night. Likewise, greater deviation in sleep onset timing was associated with greater future LEDS. Sleep onset timing appeared to have a greater magnitude of effect on future LEDS than the reverse, but this may be due to limitations in exposure measurement and/or study sample characteristics. Overall, our results suggest a bidirectional association between LEDS, a form of LAN during the sleep episode, and sleep irregularity, suggesting the importance of both maintaining a dark environment during sleep and maintaining regular sleep timing.

This study investigated individual-level light exposure during sleep (LEDS) in a naturalistic setting, which compliments the growing body of research on outdoor LAN exposure and sleep. Prior studies of satellite-measured outdoor LAN have reported associations between greater LAN and shorter sleep duration^36^, as well as greater daytime sleepiness and delayed sleep timing^37^. Similarly, studies of objective personal measures of indoor LAN have reported associations between greater LAN and worse sleep quality (LAN measured with headband device)^38,39^ and delayed sleep timing (LAN measured with light sensor placed near head)^40^. However, only limited studies examine sleep irregularity, which increasingly is linked to cardiometabolic disturbance. Phillips et al. investigated the association between LAN and the sleep regularity index in 12 regular and 12 irregular college students and reported lower daily rhythm amplitude of light exposure in irregular sleepers. While there were no differences in the daily rhythm amplitude of light exposure between the two groups, the irregular sleepers did have greater light exposure during the biological night (defined as a 10-hour window from beginning of dim light melatonin onset) when light exposure was normalized to their average daily illuminance^11^. Mead et al. analyzed repeated measures of light exposure 1-hour before sleep onset as well as light exposure during the sleep episode with sleep offset, sleep duration, sleep percentage, and sleep fragmentation index in a sample of 124 adults (light measured with wrist-worn ActiWatch) with mixed model regressions^41^. While their results did not support an association between pre-sleep light exposure with any of the sleep outcomes, a 1-unit increase in average lux during sleep (LEDS) was associated with a 0.12-hour later sleep offset timing, a 5% reduction of sleep percentage, and an increase by 0.03 of sleep fragmentation index^41^. While more research is necessary, these findings suggest LAN and LEDS may be an upstream factor contributing to sleep irregularity.

If LEDS does contribute to sleep timing regularity and disease outcomes related to circadian disruption^42^, understanding which populations have greatest exposure and why will help inform targeted sleep health interventions. In our sample, LEDS increased with age; further research is needed to investigate whether LAN or LEDS may play a role in the increased risk of developing sleep disorders and chronic health conditions among older adults, or whether chronic health conditions could lead to greater LEDS exposure. LEDS did not appear to be linked to shiftwork or insomnia in our sample, although this may differ for cohorts with higher prevalence of current employment and younger age groups. LEDS was also more prevalent among Black participants and less prevalent among Hispanic participants; this finding partly aligns with past research reporting greater outdoor neighborhood LAN exposure for Asian, Hispanic, and Black Americans compared to White Americans^43^. LEDS may also have a seasonal and geographical component, as LEDS was more prevalent in the summer or fall and less prevalent for the Minnesota study site; this may be due to greater sunlight intrusion at different times of the year and/or occlusion of the light sensor by clothing or bedsheets during colder months, as well as proximity to urban areas and nighttime light pollution. Future studies should investigate causal factors for LAN and LEDS and investigate impacts in populations which may be more sensitive to the effects, such as children, adolescents, and young adults^44–46^. The high inter-individual variability in light-induced melatonin suppression^6^ also suggests that the same LAN or LEDS may have different effects within populations.

How LEDS and variability of sleep onset timing were defined in this study should be considered when interpreting results. LEDS was defined as light during each individual’s main sleep episode, separately for each night, as a marker of light exposure during the individual’s biological night. This definition was chosen because the focus of this study was on light exposure during the biological night, rather than the solar night, and because people’s activities are shaped by their rest-wake patterns, rather than by clock time. Modeling pre-sleep light exposure may have shown larger effect estimates; however, we chose to model LEDS rather than pre-sleep light to limit possible biases of reverse causation. By focusing on the sleep episode, we may be targeting each person’s sensitive window to LAN. This contrasts with other studies of LAN that have relied on local clock time irrespective of interindividual differences or LAN during periods of low activity, such as during the 5 hours of lowest average daily activity (L5), which may miss windows where circadian phase is most sensitive to light sensitivity. Thus, how and when LAN is measured depends on how “night” is defined. LAN during the night defined by the clock on the wall and in the same way for all individuals may not appropriately reflect individual circadian phase sensitivity to light. However, these measures may be more feasible for large studies where scored sleep onset and offset information is not available. Variability of sleep onset timing was characterized in this study using person-specific measurements; each person-specific night-to-night variability was defined as the difference between two sequential starts of the main sleep episodes.

Measuring personal light exposure for large numbers of individuals and in the home environment can be challenging, and we are limited in our understanding of the light sources and reasons for LEDS exposure patterns in this study. For example, LEDS exposure (calculated as average illuminance across the sleep episode) could be due to a constant dim light source, such as a television or dim room light, or due to a brief exposure to brighter room light, such as turning on a light to use the bathroom. For example, a 5-minute exposure to 100-lux illumination during an 8-hour sleep episode (with 0 background lux) would translate to ∼1.04 average lux; the same 5-minute 100-lux exposure during an 8-hour sleep episode where the background illuminance is equal to 1 lux would translate to ∼2.03 average lux. The duration of the sleep episode would also influence the LEDS measurement. For example, a 5-minute exposure to 100-lux during a 6-hour sleep episode (with 0 background lux) would translate to ∼1.39 average lux. Both constant dim background light and/or a short pulse of bright light may plausibly influence sleep timing^47^, and future studies should consider incorporating mixed methods research to better understand the causes and sources behind LAN exposure^48^, as well as measurement of LAN/LEDS with static light sensors in the sleeping area.

There are a number of strengths and limitations for this study. Analyses were adjusted for possible confounders, including season and study site to control for seasonal-related (and latitude / climate-related) effects of light and other environment exposures on sleep, as well as physical activity, sleep fragmentation, and sleep duration. The within-individual night-to-night analysis is also a strength of the study, further supporting the logistic regression findings and testing the temporal association to provide evidence for a cause-and-effect relationship. An additional strength is the measurement of objective light and sleep patterns in a naturalistic setting of participants in their home environments. While the magnitude of effect for LEDS in this study was relatively small, this may be due to measurement limitations and/or study sample characteristics. As a wrist-worn device, the ActiWatch does not measure light exposure at the eye level, and there is measurement error when comparing it to head-worn devices^49^; however, the measurement error for LEDS (such as due to occlusion by bedsheets) would be expected to bias results towards the null^50^, although further work could simulate the direction and amount of bias given different study parameters^51^. While we evaluated light during the sleep episode as a measure of light exposure during the biological night, a marker of circadian phase (such as dim light melatonin onset) was not available. This analysis was also conducted in an older population, which may be less sensitive to the effects of inopportune light exposure due to physiological age-related changes. For example, yellowing of the eye lens with age may reduce blue-wavelength light and the amount of light intensity that reaches the retina^52,53^, suggesting that older adults may require more daytime bright light to entrain and may be less sensitive to inopportune light exposure than younger adults. Another limitation is the exclusion of participants with missing data and the potential for selection bias. Actigraphy has been shown to have some misclassification of wake and sleep episodes^54^; however, a sensitivity analysis excluding participants with insomnia did not substantially alter results.

## 5. Conclusion

Light exposure during the sleep episode (LEDS), a form of LAN, is associated with irregular sleep timing in a large sample of U.S. older adults in both averaged and night-to-night analyses, with greater light exposure predictive for larger subsequent irregularity in sleep onset timing. These results support the importance of maintaining a dark sleeping environment and maintaining regular sleep timing. Future research should aim to utilize personal measures of LAN and/or static light sensors in the sleeping area, conduct night-to-night analyses of LAN and LEDS with sleep and health outcomes, and evaluate the causes and behaviors behind LAN exposure.

## Data Availability

Actigraphy data and data from the MESA sleep ancillary study are available from the NHLBI-funded National Sleep Research Resource (NSRR): https://sleepdata.org/

## CRediT statement

**Danielle Wallace:** Conceptualization, Formal analysis, Methodology, Software, Visualization, Investigation, Writing – Original draft preparation, Writing – Reviewing and Editing, Funding acquisition. **Xinye Qiu:** Methodology, Software, conceptualization. **Joel Schwartz:** Methodology, Software, conceptualization. **Tianyi Huang:** Writing – Reviewing and Editing. **Frank Scheer:** Conceptualization, Writing – Reviewing and Editing. **Susan Redline:** Resources, Funding acquisition, Data Curation, Writing – Reviewing and Editing. **Tamar Sofer:** Methodology, Writing – Reviewing and Editing.

The views expressed in this manuscript are those of the authors and do not necessarily represent the views of the National Heart, Lung, and Blood Institute; the National Institutes of Health; or the U.S. Department of Health and Human Services. Any opinions, findings, and conclusions or recommendations expressed in this publication are those of the author(s), and do not necessarily reflect the views of the Sleep Research Society Foundation. Supported by funding from the National Institutes of Health (NIH-NHLBI T32HL007901 [to DW], K99HL166700 [to DW], R35HL135818 [to SR], R01HL098433 [to SR], R24HL114473, 75N92019R002, R01HL161012 [to TS], K01HL143034 [to TH], and R01HL155395 [to TH]). This material is also based upon work supported by the Sleep Research Society Foundation [Career Development Award to DW]. F.A.J.L.S. has been supported in part by NIH grants R01 HL140574 and R01 HL153969. MESA is conducted and supported by the National Heart, Lung, and Blood Institute in collaboration with MESA investigators. Support for MESA is provided by contracts HHSN268201500003I, N01-HC-95159, N01-HC-95160, N01-HC-95161, N01-HC-95162, N01-HC-95163, N01-HC-95164, N01-HC-95165, N01-HC-95166, N01-HC-95167, N01-HC-95168, N01-HC-95169, UL1-TR-000040, UL1-TR-001079, UL1-TR-001881, and DK06349. The MESA Sleep Exams were supported by grants from HL56984 and NIA AG070867.

## References

1. Borbély, A. A., Daan, S., Wirz-Justice, A. & Deboer, T. The two-process model of sleep regulation: a reappraisal. J. Sleep Res. 25, 131–143 (2016).

2. Czeisler, C. A. et al. Bright light induction of strong (type 0) resetting of the human circadian pacemaker. Science 244, 1328–1333 (1989).

3. Khalsa, S. B. S., Jewett, M. E., Cajochen, C. & Czeisler, C. A. A phase response curve to single bright light pulses in human subjects. J Physiol (Lond) 549, 945–952 (2003).

4. St Hilaire, M. A., et al. Human phase response curve to a 1 h pulse of bright white light. J Physiol (Lond) 590, 3035–3045 (2012).

5. Weaver, D. R. The suprachiasmatic nucleus: a 25-year retrospective. J. Biol. Rhythms 13, 100–112 (1998).

6. Phillips, A. J. K. et al. High sensitivity and interindividual variability in the response of the human circadian system to evening light. Proc Natl Acad Sci USA 116, 12019–12024 (2019).

7. Scheer, F. A. J. L. & Czeisler, C. A. Melatonin, sleep, and circadian rhythms. Sleep Med. Rev. 9, 5–9 (2005).

8. Wright, K. P. et al. Entrainment of the human circadian clock to the natural light-dark cycle. Curr. Biol. 23, 1554–1558 (2013).

9. Nordhaus, W. D. Do Real-Output and Real-Wage Measures Capture Reality?The History of Lighting Suggests Not. in The Economics of New Goods (eds. Bresnahan, T. F. & Gordon, R. J.) 29–70 (University of Chicago Press, 1996).

10. Lunn, R. M. et al. Health consequences of electric lighting practices in the modern world: A report on the National Toxicology Program’s workshop on shift work at night, artificial light at night, and circadian disruption. Sci. Total Environ. 607–608, 1073–1084 (2017).

11. Phillips, A. J. K. et al. Irregular sleep/wake patterns are associated with poorer academic performance and delayed circadian and sleep/wake timing. Sci. Rep. 7, 3216 (2017).

12. Chaput, J.-P. et al. Sleep timing, sleep consistency, and health in adults: a systematic review. Appl. Physiol. Nutr. Metab. 45, S232–S247 (2020).

13. Dzierzewski, J. M., Donovan, E. K., Kay, D. B., Sannes, T. S. & Bradbrook, K. E. Sleep inconsistency and markers of inflammation. Front. Neurol. 11, 1042 (2020).

14. Huang, T. & Redline, S. Cross-sectional and Prospective Associations of Actigraphy-Assessed Sleep Regularity With Metabolic Abnormalities: The Multi-Ethnic Study of Atherosclerosis. Diabetes Care 42, 1422–1429 (2019).

15. Yuan, R. K. et al. Chronic sleep restriction while minimizing circadian disruption does not adversely affect glucose tolerance. Front. Physiol. 12, 764737 (2021).

16. Leproult, R., Holmbäck, U. & Van Cauter, E. Circadian misalignment augments markers of insulin resistance and inflammation, independently of sleep loss. Diabetes 63, 1860–1869 (2014).

17. Mason, I. C., Qian, J., Adler, G. K. & Scheer, F. A. J. L. Impact of circadian disruption on glucose metabolism: implications for type 2 diabetes. Diabetologia 63, 462–472 (2020).

18. Chellappa, S. L., Vujovic, N., Williams, J. S. & Scheer, F. A. J. L. Impact of circadian disruption on cardiovascular function and disease. Trends Endocrinol. Metab. 30, 767–779 (2019).

19. Huang, T., Mariani, S. & Redline, S. Sleep Irregularity and Risk of Cardiovascular Events: The Multi-Ethnic Study of Atherosclerosis. J. Am. Coll. Cardiol. 75, 991–999 (2020).

20. Nikbakhtian, S. et al. Accelerometer-derived sleep onset timing and cardiovascular disease incidence: a UK Biobank cohort study. Eur. Heart J. Digit. Health 2, 658–666 (2021).

21. Münch, M. et al. The role of daylight for humans: gaps in current knowledge. Clocks & Sleep 2, 61–85 (2020).

22. Bild, D. E. et al. Multi-Ethnic Study of Atherosclerosis: objectives and design. Am. J. Epidemiol. 156, 871–881 (2002).

23. Chen, X. et al. Racial/Ethnic Differences in Sleep Disturbances: The Multi-Ethnic Study of Atherosclerosis (MESA). Sleep 38, 877–888 (2015).

24. Patel, S. R. et al. Reproducibility of a standardized actigraphy scoring algorithm for sleep in a US hispanic/latino population. Sleep 38, 1497–1503 (2015).

25. Klerman, E. B., Wang, W., Phillips, A. J. K. & Bianchi, M. T. Statistics for sleep and biological rhythms research. J. Biol. Rhythms 32, 18–25 (2017).

26. Dijk, D. J. & Czeisler, C. A. Contribution of the circadian pacemaker and the sleep homeostat to sleep propensity, sleep structure, electroencephalographic slow waves, and sleep spindle activity in humans. J. Neurosci. 15, 3526–3538 (1995).

27. Turkbey, E. B. et al. The impact of obesity on the left ventricle: the Multi-Ethnic Study of Atherosclerosis (MESA). JACC Cardiovasc. Imaging 3, 266–274 (2010).

28. Horne, J. A. & Ostberg, O. A self-assessment questionnaire to determine morningness-eveningness in human circadian rhythms. Int. J. Chronobiol. 4, 97–110 (1976).

29. Levine, D. W. et al. Reliability and validity of the Women’s Health Initiative Insomnia Rating Scale. Psychol. Assess. 15, 137–148 (2003).

30. Brown, T. M. et al. Recommendations for daytime, evening, and nighttime indoor light exposure to best support physiology, sleep, and wakefulness in healthy adults. PLoS Biol. 20, e3001571 (2022).

31. Venables, W. N. & Ripley, B. D. Modern Applied Statistics with S. (Springer, 2002).

32. Gasparrini, A. Distributed Lag Linear and Non-Linear Models in R: The Package dlnm. J. Stat. Softw. 43, 1–20 (2011).

33. Gasparrini, A., Scheipl, F., Armstrong, B. & Kenward, M. G. A penalized framework for distributed lag non-linear models. Biometrics 73, 938–948 (2017).

34. Pinheiro, J. C. & Bates, D. M. Mixed-Effects Models in S and S-PLUS. (Springer-Verlag, 2000). doi:10.1007/b98882.

35. Buysse, D. J. et al. Night-to-night sleep variability in older adults with and without chronic insomnia. Sleep Med. 11, 56–64 (2010).

36. Xiao, Q. et al. Cross-sectional association between outdoor artificial light at night and sleep duration in middle-to-older aged adults: The NIH-AARP Diet and Health Study. Environ. Res. 180, 108823 (2020).

37. Ohayon, M. M. & Milesi, C. Artificial Outdoor Nighttime Lights Associate with Altered Sleep Behavior in the American General Population. Sleep 39, 1311–1320 (2016).

38. Mitsui, K. et al. Short-wavelength light exposure at night and sleep disturbances accompanied by decreased melatonin secretion in real-life settings: a cross-sectional study of the HEIJO-KYO cohort. Sleep Med. 90, 192–198 (2022).

39. Obayashi, K., Saeki, K. & Kurumatani, N. Association between light exposure at night and insomnia in the general elderly population: the HEIJO-KYO cohort. Chronobiol. Int. 31, 976–982 (2014).

40. Chang, A.-M., Aeschbach, D., Duffy, J. F. & Czeisler, C. A. Evening use of light-emitting eReaders negatively affects sleep, circadian timing, and next-morning alertness. Proc Natl Acad Sci USA 112, 1232–1237 (2015).

41. Mead, M. P., Reid, K. J. & Knutson, K. L. Night-to-night associations between light exposure and sleep health. J. Sleep Res. 32, e13620 (2023).

42. Fishbein, A. B., Knutson, K. L. & Zee, P. C. Circadian disruption and human health. J. Clin. Invest. 131, (2021).

43. Nadybal, S. M., Collins, T. W. & Grineski, S. E. Light pollution inequities in the continental United States: A distributive environmental justice analysis. Environ. Res. 189, 109959 (2020).

44. Roenneberg, T., Allebrandt, K. V., Merrow, M. & Vetter, C. Social jetlag and obesity. Curr. Biol. 22, 939–943 (2012).

45. Zitting, K.-M. et al. Young adults are more vulnerable to chronic sleep deficiency and recurrent circadian disruption than older adults. Sci. Rep. 8, 11052 (2018).

46. Duffy, J. F., Willson, H. J., Wang, W. & Czeisler, C. A. Healthy older adults better tolerate sleep deprivation than young adults. J. Am. Geriatr. Soc. 57, 1245–1251 (2009).

47. Rahman, S. A., et al. Circadian phase resetting by a single short-duration light exposure. JCI Insight 2, e89494 (2017).

48. Sweeney, M. R. et al. Exposure to indoor light at night in relation to multiple dimensions of sleep health: Findings from the Sister Study. Sleep (2023) doi:10.1093/sleep/zsad100.

49. Jardim, A. C. N. et al. Validating the use of wrist-level light monitoring for in-hospital circadian studies. Chronobiol. Int. 28, 834–840 (2011).

50. Wallace, D. A. In the light: towards developing metrics of light regularity. Sleep (2023) doi:10.1093/sleep/zsad114.

51. Yland, J. J., Wesselink, A. K., Lash, T. L. & Fox, M. P. Misconceptions about the direction of bias from nondifferential misclassification. Am. J. Epidemiol. 191, 1485–1495 (2022).

52. Lerman, S. & Borkman, R. Spectroscopic Evaluation and Classification of the Normal, Aging, and Cataractous Lens. (With 1 color plate). Ophthalmic Res. 8, 335–353 (1976).

53. Turner, P. L. & Mainster, M. A. Circadian photoreception: ageing and the eye’s important role in systemic health. Br. J. Ophthalmol. 92, 1439–1444 (2008).

54. Sadeh, A. & Acebo, C. The role of actigraphy in sleep medicine. Sleep Med. Rev. 6, 113–124 (2002).

