## Supplemental Materials for "Light exposure during sleep is associated with irregular sleep timing: the Multi-Ethnic Study of Atherosclerosis (MESA)"

**Details on Box-Cox transformation:**

Sleep onset SD had a right-skewed distribution and was also modeled as a continuous variable after Box-Cox transformation using the “MASS” package[^31^](https://sciwheel.com/work/citation?ids=14703593&pre=&suf=&sa=0&dbf=0) using the formula (Y^λ^−1)/λ, where Y is sleep onset SD as the response variable and the transformation parameter (λ) is equal to 0.1818182. Normality of the Box-Cox transformed sleep onset SD was tested with a Shapiro-Wilk normality test (W=0.99, p=0.326).

**Details on DLNM analysis:**

Night-to-night associations were estimated with generalized additive models with penalized splines. The exposure-response function was modeled as a natural spline with internal knots selected based on smallest AIC: df=2 when LEDS or deviation in sleep onset were modeled as predictors. The lag-response function was modeled as a penalized spline with df=5 when LEDS or sleep onset modeled as predictor. Prediction variables were centered at 0. Plotting comparisons of 0 to 1 lux, 0 to 3 lux, and 0 to 10 lux were chosen to reflect a range of realistic exposure scenarios.

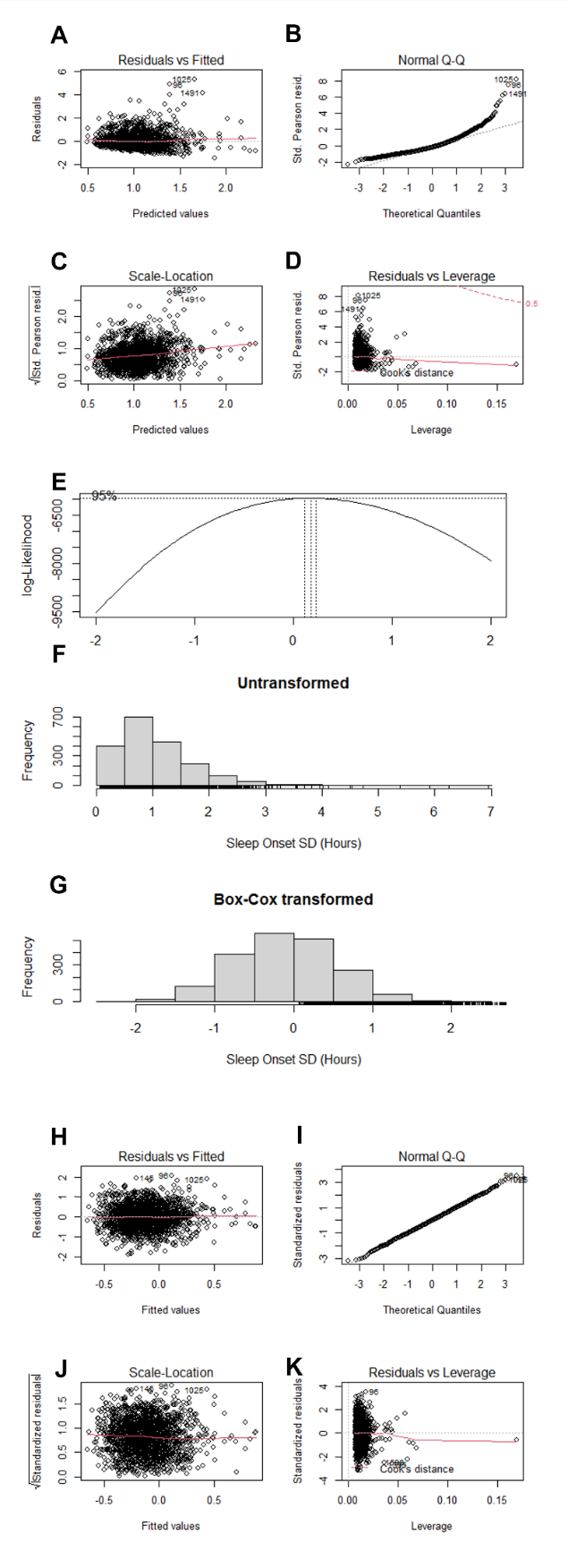

***Supplemental Figure 1****. Model fit measures and metrics comparing sleep onset SD as an untransformed versus Box-Cox transformed outcome. Model fit results of the untransformed outcome showing (****A****) a plot of the residuals versus fitted parameters, (****B****) a Q-Q plot, (****C****) a scale-location plot, and (****D****) a plot of the residuals versus leverage. A plot (****E****) of the log-likelihood profile shows the estimated lambda value, 0.18. A (****F****) histogram of the untransformed outcome shows a right-skewed distribution, compared to the (****G****) approximately normal distribution after Box-Cox transformation. The model fit results of the transformed outcome are shown with (****H****) a plot of the residuals versus fitted parameters, (****I****) a Q-Q plot, (****J****) a scale-location plot, and (****K****) a plot of the residuals versus leverage.*

| ***Supplemental Table 1****. Characteristics of the MESA Sleep study participants included (n=1,933) and not included (n=206) in the analysis.* | | | |
| --- | --- | --- | --- |
| **Variable** | **Excluded**  (n=206) | **Included**  (n=1,933) | **p-value*** |
| Age, years (mean (SD)) | 70.13 (9.70) | 69.49 (9.14) | 0.35 |
| WHR (mean (SD)) | 0.95 (0.07) | 0.94 (0.08) | 0.10 |
| Gender (N (% Male)) | 111 (53.9) | 878 (45.4) | 0.03 |
| Race/ethnicity (N (%))  *Chinese*  *Hispanic/Latino*  *Black*  *White* | 22 (10.7)  39 (18.9)  74 (35.9)  71 (34.5) | 219 (11.3)  460 (23.8)  520 (26.9)  734 (38.0) | 0.13 |
| Currently employed (N (% Yes)) | 45 (21.8) | 462 (24.0) | 0.54 |
| Income below federal poverty (N (% Yes)) | 10 (5.1) | 132 (7.1) | 0.37 |
| Partner status, married or currently living with partner (N (% Yes)) | 109 (53.7) | 1159 (60.8) | 0.06 |
| Current smoking (N (% Yes)) | 17 (8.3) | 125 (6.5) | 0.40 |
| Chronotype MEQ score (mean (SD)) | 17.00 (3.77) | 17.27 (3.61) | 0.31 |
| Season of measurement  *August-October*  *November-January*  *February-April*  *May-July* | 45 (21.8)  43 (20.9)  55 (26.7)  63 (30.6) | 505 (26.1)  433 (22.4)  486 (25.1)  509 (26.3) | 0.40 |
| Clinical Site (N (%))  *Wake Forest University*  *Columbia University*  *Johns Hopkins University*  *University of Minnesota*  *Northwestern University*  *University of California, Los Angeles* | 25 (12.1)  38 (18.4)  39 (18.9)  23 (11.2)  51 (24.8)  30 (14.6) | 308 (15.9)  343 (17.7)  283 (14.6)  364 (18.8)  336 (17.4)  299 (15.5) | 0.01 |
| **Differences in characteristics between excluded and included participants were tested using t-tests or chi-squared tests. Excluded participants are those with <6 days of valid actigraphy measures.* | | | |

| ***Supplemental Table 2****. Distribution of the Night 5 to Night 6 sleep deviation values and Night 5 LEDS values for outcomes in the night-to-night analysis.* | | | | | |
| --- | --- | --- | --- | --- | --- |
| **Sample distribution of variation in sleep onset (SD across days)** | | | | | |
| **<1 hour** | **1-2 hours** | **2-3 hours** | **3-4 hours** | **4-5 hours** | **5+ hours** |
| N=1098 (57%) | N=662 (34.2%) | N=146 (7.6%) | N=17 (0.9%) | N=6 (0.3%) | N=4 (0.2%) |
| **Sample distribution of \|Night 5 – Night 6\| sleep deviation values** | | | | | |
| **<1 hour** | **1-2 hours** | **2-3 hours** | **3-4 hours** | **4-5 hours** | **5+ hours** |
| N=1087 (56%) | N=473 (24.5%) | N=179 (9.3%) | N=96 (5.0%) | N=46 (2.4%) | N=52 (2.7%) |
| **Sample distribution of averaged (across days) LEDS lux values** | | | | | |
| **0-0.5 lux** | **0.5-1 lux** | **1-2 lux** | **2-3 lux** | **3-10 lux** | **10+ lux** |
| N=878 (45.4%) | N=446 (23.1%) | N=275 (14.2%) | N=81 (4.2%) | N=178 (9.2%) | N=75 (3.9%) |
| **Sample distribution of Night 5 LEDS lux values** | | | | | |
| **0-0.5 lux** | **0.5-1 lux** | **1-2 lux** | **2-3 lux** | **3-10 lux** | **10+ lux** |
| N=1124 (58.7%) | N=340 (17.8%) | N=173 (9.0%) | N=67 (3.5%) | N=137 (7.2%) | N=73 (3.8%) |

| ***Supplemental Table 3****. Linear regression results for light exposure during sleep (LEDS, continuous or tertiles) with Box-Cox-transformed sleep onset timing variability (SD, hours) as the outcome.* | | | | | | |
| --- | --- | --- | --- | --- | --- | --- |
|  | Crude  [95% CI] | Model 1  [95% CI] | Model 2  [95% CI] | Model 3  [95% CI] | **Model 4**  **[95% CI]** | Model 5  [95% CI] |
| *LEDS as continuous*:* | | | | | | |
| *LEDS (per 1-unit lux)* | **0.02**  (0.02-0.03) | **0.02**  (0.01-0.03) | **0.02**  (0.01-0.03) | **0.02**  (0.01-0.03) | **0.02**  (0.01-0.03) | **0.02**  (0.01-0.03) |
| *LEDS as tertiles*:* | | | | | | |
| *LEDS T1 (0-0.33 lux)* | *(ref)* | *(ref)* | *(ref)* | *(ref)* | *(ref)* | *(ref)* |
| *LEDS T2 (0.33-0.95 lux)* | 0.03  (-0.04-0.09) | 0.01  (-0.06-0.07) | 0.00  (-0.06-0.07) | 0.01  (-0.06-0.08) | 0.02  (-0.05-0.08) | 0.02  (-0.05-0.08) |
| *LEDS T3 (0.95-39.4 lux)* | **0.22**  (0.16-0.29) | **0.21**  (0.14-0.28) | **0.21**  (0.14-0.28) | **0.20**  (0.13-0.27) | **0.20**  (0.13-0.27) | **0.20**  (0.13-0.27) |
| **Sleep onset SD (hours) as outcome*  *Crude and adjusted model regression estimates with 95% confidence intervals are presented, with p<0.05 in bold. Model 1 adjusted for age, gender, and race/ethnicity; Model 2 adjusted for all the covariates included in Model 1 in addition to poverty, employment status, and partner status; Model 3 adjusted for all the covariates included in Model 2 in addition to smoking, chronotype, waist to hip ratio, and average behavioral activity; Model 4 adjusted for all the covariates included in Model 3 in addition to season and Exam 5 site; Model 5 adjusted for all the covariates in Model 4 in addition to average sleep episode duration, and average sleep fragmentation index.* | | | | | | |

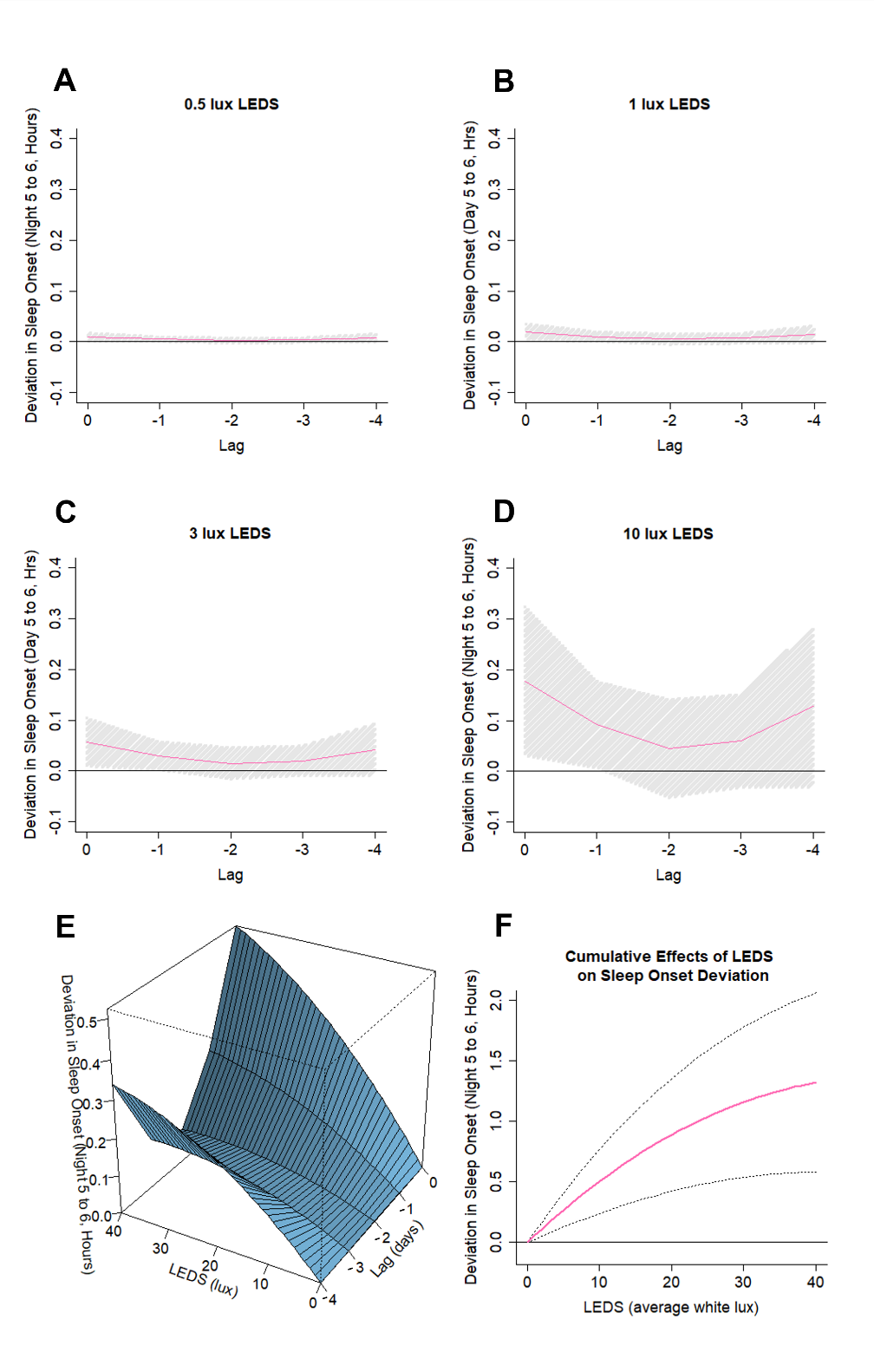

***Supplemental Figure 2.*** *Plots of predicted lagged effects of LEDS (5^th^ night=lag0, 4^th^ night=lag-1, 3^rd^ night=lag-2, 2^nd^ night=lag-3, and 1^st^ night=lag-4) on sleep onset deviation (the absolute difference in sleep onset timing between the 5^th^ and 6^th^ night). The analysis was adjusted for age, gender, and race/ethnicity. Figures showing: the predicted lagged effects of (****A****) 0.5 lux LEDS exposure on sleep onset deviation, (****B****) 1 lux LEDS exposure on sleep onset deviation, (****C****) 3 lux LEDS exposure on sleep onset deviation, and (****D****) 10 lux LEDS exposure on sleep onset deviation. Plots showing the (****E****) overall 3-dimensional exposure-outcome cross-basis matrix and the (****F****) cumulative lagged effects of LEDS on sleep onset deviation.*

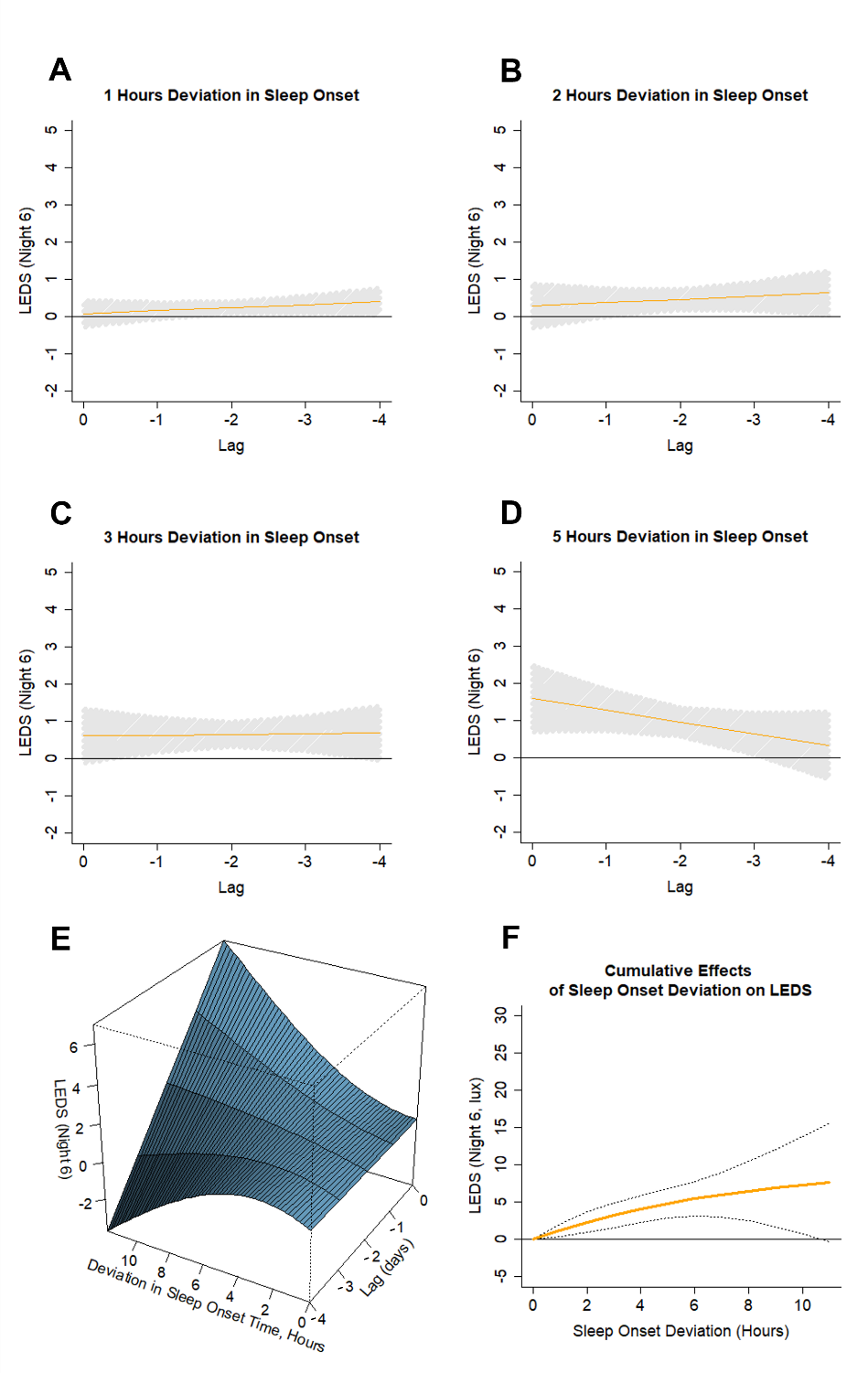

***Supplemental Figure 3.*** *Plots of predicted lagged effects of sleep onset deviation (absolute difference in sleep onset timing between the 5^th^ and 6^th^ night=lag0, 4^th^ and 5^th^ night=lag-1, between the 3^rd^ and 4^th^ night=lag-2, between the 2^nd^ and 3^rd^ night=lag-3, between the 1^st^ and 2^nd^ night=lag-4) on LEDS lux during the 6^th^ night. The analysis was adjusted for age, gender, and race/ethnicity. Figures showing the predicted lagged effects of (****A****) 1 hour sleep onset deviation on LEDS, (****B****) 2 hours sleep onset deviation on LEDS, (****C****) 3 hours sleep onset deviation on LEDS, and (****D****) 5 hours sleep onset deviation on LEDS. Plots showing the (****E****) overall 3-dimensional exposure-outcome cross-basis matrix and the (****F****) cumulative lagged effects of sleep onset deviation on LEDS.*

| ***Supplemental Table 4****. Logistic regression sensitivity analysis results excluding shift workers (n=235) for light exposure during sleep (LEDS, continuous or tertiles) with irregular sleep onset (≥1.36 hours SD) as the outcome.* | | | | | | |
| --- | --- | --- | --- | --- | --- | --- |
|  | Crude  [95% CI] | Model 1  [95% CI] | Model 2  [95% CI] | Model 3  [95% CI] | **Model 4**  **[95% CI]** | Model 5  [95% CI] |
| *LEDS as continuous*:* | | | | | | |
| *LEDS (per 1-lux unit)* | **1.08**  (1.05-1.11) | **1.08**  (1.05-1.11) | **1.08**  (1.05-1.11) | **1.07**  (1.04-1.10) | **1.07**  (1.04-1.10) | **1.06**  (1.03-1.09) |
| *LEDS as tertiles*:* | | | | | | |
| *LEDS T1* | *(ref)* | *(ref)* | *(ref)* | *(ref)* | *(ref)* | *(ref)* |
| *LEDS T2* | 1.04  (0.78-1.39) | 0.98  (0.73-1.31) | 0.95  (0.7-1.29) | 0.99  (0.73-1.35) | 0.99  (0.72-1.34) | 1.03  (0.75-1.42) |
| *LEDS T3* | **1.84**  (1.40-2.43) | **1.75**  (1.32-2.33) | **1.75**  (1.31-2.34) | **1.76**  1.31-2.38) | **1.72**  (1.27-2.34) | **1.64**  (1.19-2.26) |
| *Crude and adjusted odds ratios (ORs) with 95% confidence intervals are presented. ORs with p<0.05 are in bold. Model 1 adjusted for age, gender, and race/ethnicity; Model 2 adjusted for all the covariates included in Model 1 in addition to poverty, employment status, and partner status; Model 3 adjusted for all the covariates included in Model 2 in addition to smoking, chronotype, waist to hip ratio, and average behavioral activity; Model 4 adjusted for all the covariates included in Model 3 in addition to season and Exam 5 site; Model 5 adjusted for all the covariates in Model 4 in addition to average sleep episode duration and average sleep fragmentation index.* | | | | | | |

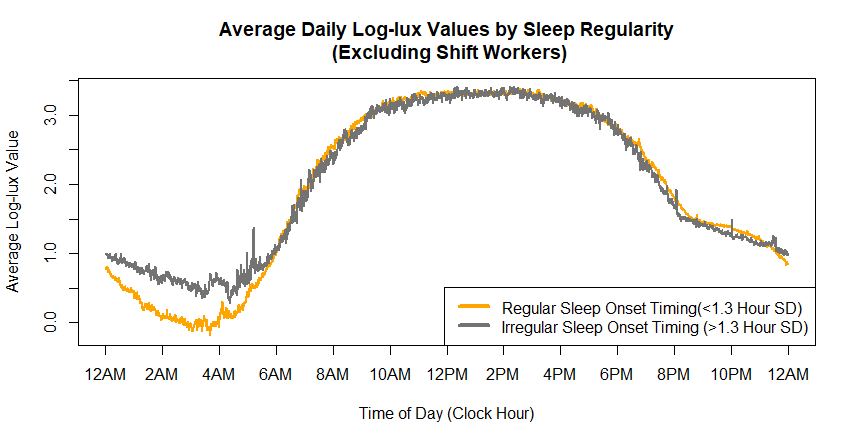

***Supplemental Figure 4****. Average daily log10-lux illuminance values in MESA by time of day, excluding shift workers (n=235), stratified by the regular sleep onset timing group (n=1,311 participants) and the irregular sleep onset timing group (n=387 participants).*

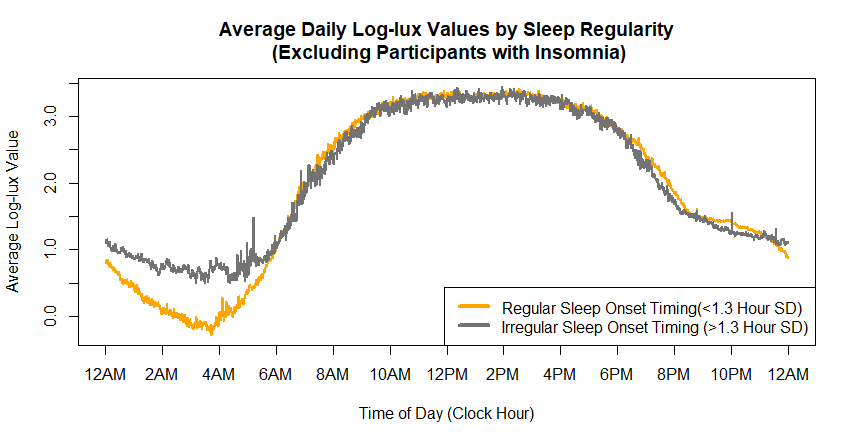

***Supplemental Figure 5****. Average nightly log10-lux illuminance values in MESA by time of day, excluding participants with insomnia (n=717), stratified by the regular sleep onset timing group (n=943 participants) and the irregular sleep onset timing group (n=273 participants).*

| ***Supplemental Table 5.*** *Sensitivity analysis excluding shift workers (n=235) for mixed model regression of night-to-night associations between LEDS (exposure) and absolute deviation in sleep onset (outcome).* | | | | | |
| --- | --- | --- | --- | --- | --- |
|  |  | **Deviation in Sleep Onset (hours)** | | | |
|  | **LEDS** | β | SE | t | p-value |
| Crude | 1-lux unit | 0.008 | 0.002 | 4.039 | 5.42E-05 |
| Model 1 | 1-lux unit | 0.008 | 0.002 | 4.060 | 4.97E-05 |
| Model 2 | 1-lux unit | 0.008 | 0.002 | 4.086 | 4.44E-05 |
| Model 3 | 1-lux unit | 0.007 | 0.002 | 3.918 | 9.02E-05 |
| **Model 4** | 1-lux unit | 0.007 | 0.002 | 3.820 | 1.35E-04 |
| Model 5 | 1-lux unit | 0.008 | 0.002 | 4.171 | 3.07E-05 |
| *Results from mixed model regression. Model 1 adjusted for age, gender, and race/ethnicity; Model 2 adjusted for all the covariates included in Model 1 in addition to poverty, employment status, and partner status; Model 3 adjusted for all the covariates included in Model 2 in addition to smoking, chronotype, waist to hip ratio, and average behavioral activity; Model 4 adjusted for all the covariates included in Model 3 in addition to season and Exam 5 site; Model 5 adjusted for all the covariates in Model 4 in addition to nightly sleep episode duration and nightly sleep fragmentation index.* | | | | | |

| ***Supplemental Table 6****. Logistic regression sensitivity analysis results excluding participants with insomnia (n=717) for light exposure during sleep (LEDS, continuous or tertiles) with irregular sleep onset (≥1.36 hours SD) as the outcome.* | | | | | | |
| --- | --- | --- | --- | --- | --- | --- |
|  | Crude  [95% CI] | Model 1  [95% CI] | Model 2  [95% CI] | Model 3  [95% CI] | **Model 4**  **[95% CI]** | Model 5  [95% CI] |
| *LEDS as continuous*:* | | | | | | |
| *LEDS (per 1-unit lux)* | **1.09**  (1.05-1.12) | **1.09**  (1.06-1.12) | **1.09**  (1.06-1.13) | **1.09**  (1.05-1.12) | **1.09**  (1.05-1.12) | **1.08**  (1.04-1.12) |
| *LEDS as tertiles*:* | | | | | | |
| *LEDS T1* | *(ref)* | *(ref)* | *(ref)* | *(ref)* | *(ref)* | *(ref)* |
| *LEDS T2* | 1.25  (0.89-1.77) | 1.18  (0.83-1.68) | 1.08  (0.75-1.56) | 1.16  (0.80-1.68) | 1.14  (0.78-1.66) | 1.23  (0.83-1.83) |
| *LEDS T3* | **2.21**  (1.58-3.09) | **2.11**  (1.5-2.99) | **2.14**  (1.51-3.05) | **2.20**  (1.53-3.17) | **2.16**  (1.50-3.14) | **2.07**  (1.41-3.06) |
| *Crude and adjusted odds ratios (ORs) with 95% confidence intervals are presented. ORs with p<0.05 are in bold. Model 1 adjusted for age, gender, and race/ethnicity; Model 2 adjusted for all the covariates included in Model 1 in addition to poverty, employment status, and partner status; Model 3 adjusted for all the covariates included in Model 2 in addition to smoking, chronotype, waist to hip ratio, and average behavioral activity; Model 4 adjusted for all the covariates included in Model 3 in addition to season and Exam 5 site; Model 5 adjusted for all the covariates in Model 4 in addition to average sleep episode duration and average sleep fragmentation index.* | | | | | | |

| ***Supplemental Table 7.*** *Sensitivity analysis excluding participants with insomnia (n=717) for mixed model regression of night-to-night associations between LEDS and absolute deviation in sleep onset.* | | | | | |
| --- | --- | --- | --- | --- | --- |
|  |  | **Deviation in Sleep Onset (hours)** | | | |
|  | **LEDS** | β | SE | t | p-value |
| Crude | 1-lux unit | 0.006 | 0.002 | 2.886 | 0.004 |
| Model 1 | 1-lux unit | 0.006 | 0.002 | 2.968 | 0.003 |
| Model 2 | 1-lux unit | 0.006 | 0.002 | 2.971 | 0.003 |
| Model 3 | 1-lux unit | 0.006 | 0.002 | 2.781 | 0.005 |
| **Model 4** | 1-lux unit | 0.006 | 0.002 | 2.754 | 0.006 |
| Model 5 | 1-lux unit | 0.006 | 0.002 | 2.888 | 0.004 |
| *Results from mixed model regression. Model 1 adjusted for age, gender, and race/ethnicity; Model 2 adjusted for all the covariates included in Model 1 in addition to poverty, employment status, and partner status; Model 3 adjusted for all the covariates included in Model 2 in addition to smoking, chronotype, waist to hip ratio, and average behavioral activity; Model 4 adjusted for all the covariates included in Model 3 in addition to season and Exam 5 site; Model 5 adjusted for all the covariates in Model 4 in addition to nightly sleep episode duration and nightly sleep fragmentation index.* | | | | | |

| ***Supplemental Table 8.*** *Sensitivity analysis excluding shift workers (n=235) for mixed model regression for night-to-night associations between absolute deviation in sleep onset (exposure) and LEDS (outcome).* | | | | | |
| --- | --- | --- | --- | --- | --- |
|  |  | **LEDS (lux)** | | | |
|  | **Deviation** | β | SE | t | p-value |
| Crude | 1-hour units | 0.218 | 0.068 | 3.197 | 0.001 |
| Model 1 | 1-hour units | 0.219 | 0.068 | 3.209 | 0.001 |
| Model 2 | 1-hour units | 0.220 | 0.068 | 3.221 | 0.001 |
| Model 3 | 1-hour units | 0.208 | 0.068 | 3.039 | 0.002 |
| **Model 4** | 1-hour units | 0.203 | 0.068 | 2.966 | 0.003 |
| Model 5 | 1-hour units | 0.203 | 0.068 | 2.973 | 0.003 |
| *Results from mixed model regression. Model 1 adjusted for age, gender, and race/ethnicity; Model 2 adjusted for all the covariates included in Model 1 in addition to poverty, employment status, and partner status; Model 3 adjusted for all the covariates included in Model 2 in addition to smoking, chronotype, waist to hip ratio, and average behavioral activity; Model 4 adjusted for all the covariates included in Model 3 in addition to season and Exam 5 site; Model 5 adjusted for all the covariates in Model 4 in addition to nightly sleep episode duration and nightly sleep fragmentation index.* | | | | | |

| ***Supplemental Table 9.*** *Sensitivity analysis excluding participants with insomnia (n=717) for mixed model regression for night-to-night associations between absolute deviation in sleep onset (exposure) and LEDS (outcome).* | | | | | |
| --- | --- | --- | --- | --- | --- |
|  |  | **LEDS (lux)** | | | |
|  | **Deviation** | β | SE | t | p-value |
| Crude | 1-hour units | 0.560 | 0.087 | 6.404 | 1.67E-10 |
| Model 1 | 1-hour units | 0.562 | 0.088 | 6.375 | 2.01E-10 |
| Model 2 | 1-hour units | 0.561 | 0.088 | 6.359 | 2.23E-10 |
| Model 3 | 1-hour units | 0.545 | 0.088 | 6.173 | 7.30E-10 |
| **Model 4** | 1-hour units | 0.544 | 0.088 | 6.156 | 8.09E-10 |
| Model 5 | 1-hour units | 0.545 | 0.088 | 6.161 | 7.84E-10 |
| *Results from mixed model regression. Model 1 adjusted for age, gender, and race/ethnicity; Model 2 adjusted for all the covariates included in Model 1 in addition to poverty, employment status, and partner status; Model 3 adjusted for all the covariates included in Model 2 in addition to smoking, chronotype, waist to hip ratio, and average behavioral activity; Model 4 adjusted for all the covariates included in Model 3 in addition to season and Exam 5 site; Model 5 adjusted for all the covariates in Model 4 in addition to nightly sleep episode duration and nightly sleep fragmentation index.* | | | | | |
